# DIETARY MANIPULATION OF THE GUT MICROBIOME IN INFLAMMATORY BOWEL DISEASE PATIENTS: PROOF OF CONCEPT

**DOI:** 10.1101/2021.10.07.21250296

**Authors:** Barbara Olendzki, Vanni Bucci, Caitlin Cawley, Rene Maserati, Margaret McManus, Effie Olendzki, Camilla Madziar, David Chiang, Doyle V. Ward, Randall Pellish, Christine Foley, Shakti Bhattarai, Beth A. McCormick, Ana Maldonado-Contreras

## Abstract

Diet is a modifiable, non-invasive, inexpensive behavior that is crucial in shaping the intestinal microbiome. A microbiome “imbalance” or dysbiosis in inflammatory bowel disease (IBD) is linked to inflammation. Here, we aim to define the impact of specific foods on bacterial species commonly depleted in patients with IBD to better inform dietary treatment. We performed a single-arm, pre-post intervention trial. After a baseline period, a dietary intervention with the IBD-Anti-Inflammatory Diet (IBD-AID) was initiated. We collected stool and blood samples and assessed dietary intake throughout the study. We applied advanced computational approaches to define and model complex interactions between the foods reported and the microbiome. A dense dataset comprising 553 dietary records and 340 stool samples was obtained from 22 participants. Consumption of prebiotics, probiotics, and beneficial foods correlated with increased abundance of *Clostridia* and *Bacteroides*, commonly depleted in IBD cohorts. We further show that the IBD-AID intervention affects the immune tone by lowering IL-8 and increasing GM-CSF with certain foods correlating with levels of those cytokines. By using robust predictive analytics, this study represents the first steps to detangle diet-microbiome interactions to inform personalized nutrition for patients suffering from dysbiosis-related IBD.

## INTRODUCTION

The etiology of Inflammatory bowel disease (IBD) is thought to be linked to an inappropriate immune response to an altered, dysbiotic gut microbiome. Dysbiosis in IBD patients is characterized by depletion of *Clostridia* and *Bacteroides* (1-6). These bacterial species are known to maintain gut homeostasis via the production of short-chain fatty acids (SCFAs) (7-11). Dietary interventions represent an ideal strategy to revert gut dysbiosis in IBD patients as diet change is often more embraced by individuals than medication (12). Also, diet is safe, does not require FDA approval (12), and has been proven to rapidly change the microbiome (13).

Recent trials have demonstrated that dietary therapy is effective for pediatric patients with Crohn’s disease. The diets tested as a therapy for pediatric patients included the Specific Carbohydrate Diet (SCD), the modified SCD (mSCD, which includes oats), the Crohn’s disease exclusion diet with partial enteral nutrition, and the exclusive enteral nutrition diet (14-16). In those studies, more than 80% of patients achieved clinical remission between 4- and 6-weeks post-intervention. Diet favored increased abundance of *Clostridia* species, including *Faecalibacterium prausnitzii, Roseburia hominis*, and *Eubacterium eligens* (14-16).

In adults with Crohn’s disease, a recent randomized trial that included interventions with either the SCD or the Mediterranean diet has also demonstrated a remarkable effect of diet in inducing remission (17). Specifically, after only 6 weeks on either diet half of the patients in the trial achieved symptomatic remission with >30% showing reduction of fecal calprotectin levels (17). For ulcerative colitis, a catered nutritious low-fat/high-fiber diet has been shown to improve the overall quality of life, lower inflammatory markers, decreased dysbiosis, and specifically favor *Faecalibacterium prausnitzi* (18). We also designed the IBD-Anti-Inflammatory Diet or IBD-AID (19, 20). In a retrospective study, we reported that adult patients, both Crohn’s disease or ulcerative colitis patients, adopting the IBD-AID experienced reduction of disease activity and lowered their medication intake only after 4 weeks on the diet (20). The IBD-AID has been designed to revert dysbiosis in patients with IBD.

In this current work, we sought to rigorously establish whether adherence to the IBD-AID can revert dysbiosis by favoring SCFA-producing bacteria that are depleted in patients with IBD. We leveraged our robust and validated predictive analytic and mathematical modeling (21-23) to perform fine-scale analysis of bacterial species favored by specific foods during an 8-week dietary intervention with the IBD-AID.

## RESULTS

### Demographics of the participants of the study

We enrolled 25 subjects with CD or UC to complete an 8-week IBD-AID dietary intervention **(Figure 1A)**. A total of 22 participants completed the baseline period (age average = 40.5 ± 12.8. **Table 1**). Nineteen subjects continued to complete the intervention period (12 CD and 7 UC). The average body mass index (BMI) for participants in the study was 27.9 ± 5.8 (overweight and obese), which is comparable to the average BMI among Americans (24). Only 1 UC participant was underweight (BMI = 17.9). Except for 2 CD participants reporting no IBD-related medications, participants were using biologics (31.8%), aminoacylates (27.2%), steroids (22.7%), and immunomodulators (13.60%).

**Figure 1.**
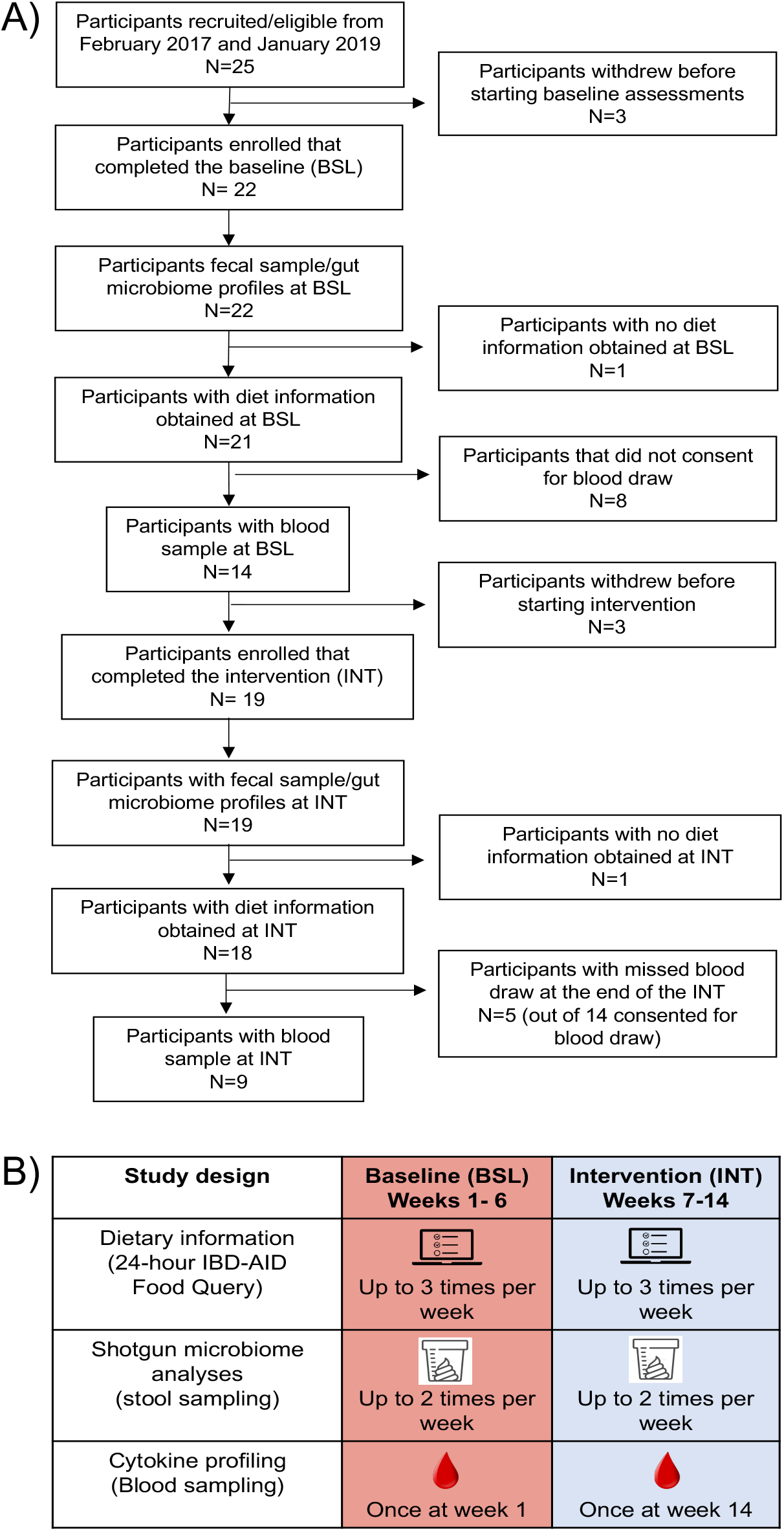
A) Participant inclusion and exclusion during the study duration. B) A schematic representation of the study design which involved bi-weekly stool samples collection and completion of 24-hour IBD-AID Food Querys up to three times a week throughout the study. At the beginning of the baseline and the end of the intervention, blood samples were collected.

**Table 1.** Demographic description of all the participants recruited for the study between February 2017 and January 2019.

| Patient information | Participants included in analyses (n=22) |  | Enrolled participants (n=25) |  |
| --- | --- | --- | --- | --- |
|  | Crohn's disease (n=15) | Ulcerative colitis (n=7) | Crohn's disease (n=16) | Ulcerative colitis (n=9) |
| <u>Demographics</u> |  |  |  |  |
| Average age (years) | 41.7 ± 13.4 | 37.8 ± 11.5 | 41.4 ± 12.9 | 39.6 ± 11.5 |
| Average weight (lbs) | 173.6 ± 31.4 | 190.1 ± 57.0 | 171.5 ± 31.3 | 190.1 ± 57.0 |
| Average BMI | 29.3 ± 5.4 | 25.4 ± 6.7 | 29.5 ± 5.2 | 25.4 ± 6.7 |
| Female sex (%) | 11 (73.3%) | 2 (28.5%) | 12 (75%) | 3 (33%) |
| White race (%) | 14 (93.3%) | 6 (85.7%) | 14 (87.5%) | 8 (88.8%) |
| <u>IBD Medications</u> |  |  |  |  |
| Current use of Amisosalicylates | 3 (20%) | 3 (42.8%) | 3 (18.7%) | 4 (44.4%) |
| Current use of Biologics | 6 (40%) | 1 (14.2%) | 6 (37.5%) | 1 (11.1%) |
| Current use of Immnomodulators | 1 (6.6%) | 2 (28.5%) | 1 (6.2%) | 2 (22.2%) |
| Current use of Steroids | 3 (20%) | 2 (28.5%) | 4 (25%) | 3 (33%) |
| <u>Other Medications</u> |  |  |  |  |
| Current use of Antihistamine | 4 (26.6%) | 0 | 5 (31.2%) | 0 |
| Current use of SSRI | 4 (26.6%) | 2 (28.5%) | 4 (25%) | 3 (33%) |
| Current use of Diuretic | 4 (26.6%) | 0 | 5 (31.2%) | 0 |
| Current use of Vitamin D supplement | 4 (26.6%) | 2 (28.5%) | 4 (25%) | 4 (44.4%) |

### Subjects profoundly changed their diet during the intervention

At baseline, we obtained 134 and 89 unique 24-hour IBD-AID Food Querys from 14 CD and 7 UC participants, respectively. We observed that all the participants reported similar diets at baseline (Mann-Whitney test, p-value > 0.5. **Supplementary Table 2**), except for intakes of lean animal protein (included in beneficial foods), which was higher in UC patients. As expected, participants reported a low intake of fruits and vegetables comparable to an average American (25).

At the intervention, we obtained 218 and 112 unique 24-hour IBD-AID Food Querys from 11 CD and 7 UC participants, respectively. We observed that overall, participants profoundly changed their diet reporting an average of 1.8-fold increase in prebiotics consumption, a 1.5-fold increase in probiotics consumption, a 1.6-fold increase in beneficial foods consumption, and a 3.7-fold reduction in adverse foods consumption **(Figure 2A. Table 2)**. More detailed analyses showed that participants significantly increased their intake of all foods contained in the prebiotic category, fermented dairy products within probiotic foods, and omega 3 fatty acids from the beneficial foods category. In contrast, participants significantly reduced consumption of most of the foods included in the adverse food category with exception of artificial sweeteners (**Figure 2B**). We also observed that changes in food intake occurred within the first weeks of the intervention (**Figure 2C**), suggesting rapid adaptation to the diet.

**Figure 2.**
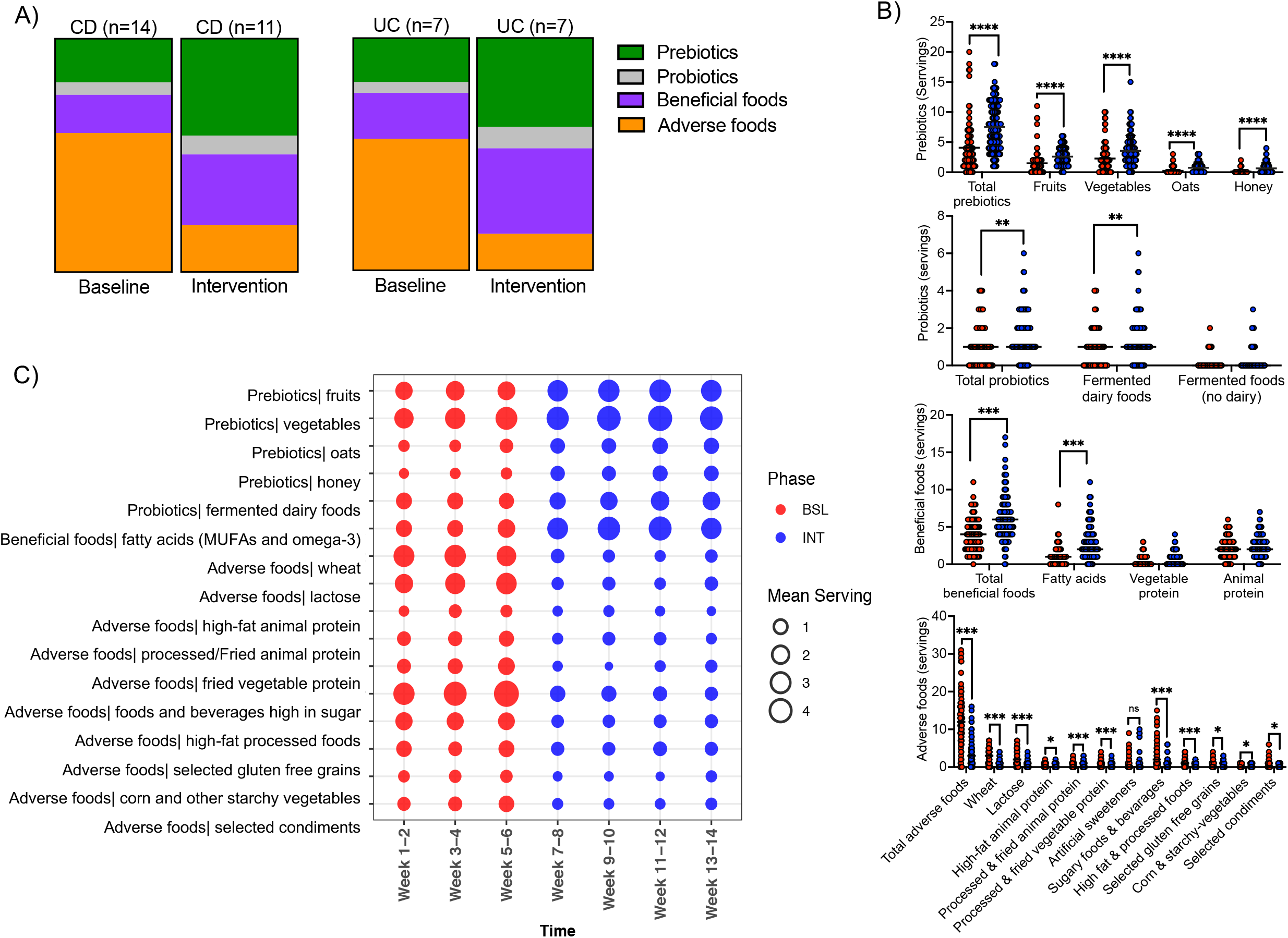
Participants adhere to the IBD-AID. A) Average serving size of foods categories consumed at baseline (BSL) and intervention (INT). B) Food categories with increased consumption during the intervention (Multiple T-test, p-value < 0.05). The mean intakes per study period: BSL or INT, was calculated on the average intake per food category per week. C) Reported servings of prebiotics, probiotics, beneficial foods, and adverse foods per week at BSL (in red) and INT (in blue) period. Each circle represents the mean intake per food category grouped in 2 weeks intervals.

**Table 2.** Mean servings reported on the 24-hour IBD-AID Food Query at baseline and intervention.

| Food categories | Mean servings/d reported at BSL | Mean servings/d reported at INT | Difference between means (BSL - INT) ± SEM |
| --- | --- | --- | --- |
| Prebiotics | 4.08 | 7.51 | 3.44 ± 0.58 |
| Probiotics | 1.09 | 1.59 | 0.50 ± 0.17 |
| Beneficial foods | 3.88 | 6.13 | 2.26 ± 0.37 |
| Adverse foods | 12.64 | 3.42 | -9.23 ± 0.86 |

Separating by disease phenotype, we observed that CD and UC participants reported a similar increase in intake of foods encouraged during the intervention, except for oats and vegetable protein, which were only significantly increased in UC or CD participants, respectively. Intakes of processed fried animal protein, corn, and starchy vegetables were only significantly decreased on CD participants; and selected avoided condiments (i.e., wheat-based soy sauce, Sriracha, ketchup, relish, BBQ sauce) only decreased in UC participants (**Supplementary Tables 3 and 4**). CD participants reported no consumption of artificial sweeteners while UC participants did consume this food item. This might explain the lack of differences described above.

Lastly, during the intervention alcohol consumption was reported higher for CD participants (Mann-Whitney test, p-value < 0.01. **Supplementary Table 3)**. However, there were no differences in alcohol consumption between study periods. In UC participants, there was a trend of decreasing alcohol consumption during the intervention (Mann-Whitney test, p-value = 0.1), which might explain the differences in alcohol intakes between the CD and UC participants.

In sum, we observed that overall participants can rapidly adopt the IBD-AID.

### The IBD-AID favors SCFA-producing bacterial species

We collected a total of 340 stool samples: 143 at baseline and 197 during the intervention. The average number of stool samples per participant was 6.5 ± 2.1 at baseline (n=22) and 10.3 ± 5.1 at intervention (n=19). We observe high microbiome inter-personal variability among participants with no differences by disease phenotype (CD vs UC) in alpha and beta diversity (**Supplementary Figure 1)** nor in microbiota representation at baseline (BH p-value > 0.05, data not shown).

We then investigated the impact of the intervention on the gut microbiome. First, we did not find differences in alpha and beta diversity between samples collected at baseline *vs*. intervention (**Supplementary Figure 2**). However, compared to baseline, we found specific bacterial species have a reduced or increased abundance during the intervention window (BH-adjusted p-value < 0.05). The top 10 bacteria with increased abundance in both CD and UC participants during intervention are SCFA-producing bacteria mostly belonging to the *Clostridia* class **(Figure 3A**). Overall, the increased abundance of *Roseburia hominis* distinguished the highest likelihood samples collected during the intervention. Conversely, reduced abundance of members of the *Bacteroidia, Coriobacteriia, Clostridia*, and *Negativicutes* classes predicted the highest likelihood samples collected during the intervention (**Figure 3B**).

**Figure 3.**
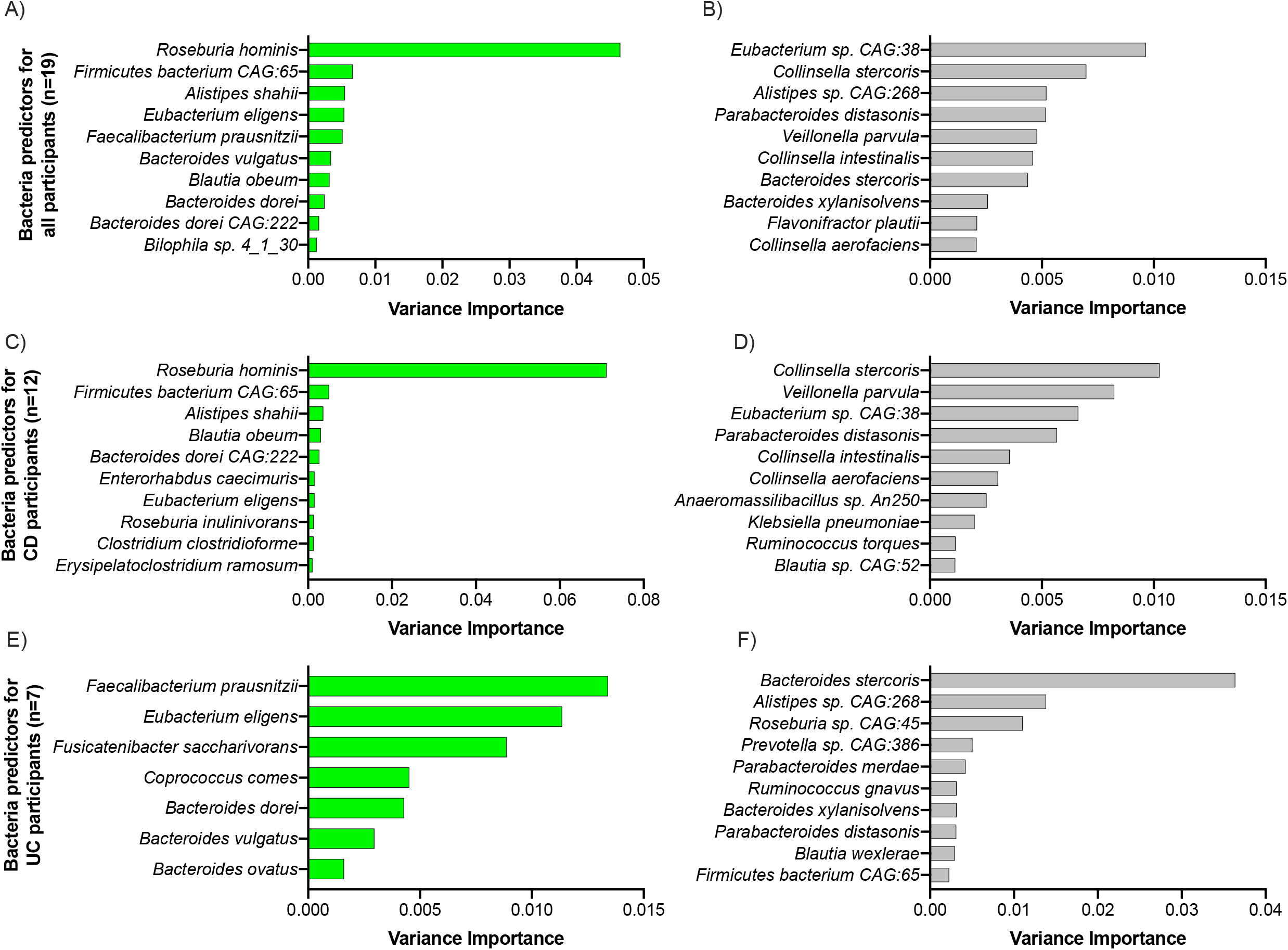
Mixed effect random forest classification analysis identified microbes affected by the intervention. Bar plots show the variance of the importance of bacterial species found to be enriched (in green) or depleted (in gray) during the intervention in all (A and B), CD (C and D), and UC (E and F) subjects completing the intervention (BH p-value > 0.05).

We then investigated whether different species could be enriched by disease phenotype (**Supplementary Table 5**). In patients with CD, the top bacteria with significantly increased abundance during the intervention were mostly species members of the *Clostridia, Bacteroidia*, and *Coriobacteriia* classes, and 2 *Firmicutes* species (**Figure 3C**). Bacterial species significantly reduced during the intervention belonged not only to *Gammaproteobacteria* and Negativicutes classes, but also to *Clostridia, Bacteroidia*, and *Coriobacteriia* classes (**Figure 3D**). Despite the overlap of bacterial classes as being positively or negatively affected by the IBD-AID, there were specific species within those classes that seemed to be directionally altered by the intervention. These results suggest that specific foods affect the abundance of bacteria at the species level and are consistent with previous studies (26-28).

In subjects with UC, similar results were observed. The abundance of specific *Clostridia* and *Bacteroides* species known to be depleted in UC patients (i.e., *Eubacterium eligens, Faecalibacterium prausnitzii, Fusicatenibacter saccharivorans, Bacteroides dorei, Bacteroides ovatus*, and *Bacteroides vulgatus)* was significantly increased during the intervention. Conversely, other *Clostridia* and *Bacteroides* were significantly decreased at intervention **(Figure 3E and 3F)**.

Taken together, these findings show an overall shift of the microbiome after the intervention that differs by disease phenotype and is specie specific. The top bacteria favored by the intervention were *Roseburia hominis* and *Faecalibacterium prautnizii* in CD and UC subjects, respectively. Only 2 species, *Eubacterium eligens*, and *Bacteroides dorei* were enriched in both CD and UC, while *Parabacteroides distasonis* was consistently decreased in all participants regardless of disease phenotype during the intervention.

### The IBD-AID favors a microbiome with anti-inflammatory capacity

We next evaluated the functional capacity of the microbiome after the intervention. At baseline, we found that the metagenomic capacity varied greatly by participant, with most of the clustering of the samples by participant (data not shown). However, we observe that during the intervention the microbiome exhibited an increased genetic capacity for 1) biosynthesis of several key amino acids (i.e., histidine, lysine, threonine, methionine, serine, glycine, isoleucine, and arginine); 2) degradation of mannan (a dietary fiber); and 3) β-oxidation for fatty acid degradation (**Figure 4A**). *Roseburia* sp. and *Faecalibacterium* sp. – both favored during the IBD-AID intervention are main degraders of dietary mannan ultimately producing SCFA (29, 30). Mannans are found in the endospermic tissue of nuts (homopolymeric mannan), barley, oats (β-glucans or mannoproteins), coffee beans, coconut palm, tomato, and legume seeds (galactomannan) (31). Similarly, increased microbiome gene capacity for oxidation of fatty acids also suggests increased availability of SCFAs. Thus, we further investigated the impact of IBD-AID on the pool of microbial genes involved in SCFA production.

**Figure 4.**
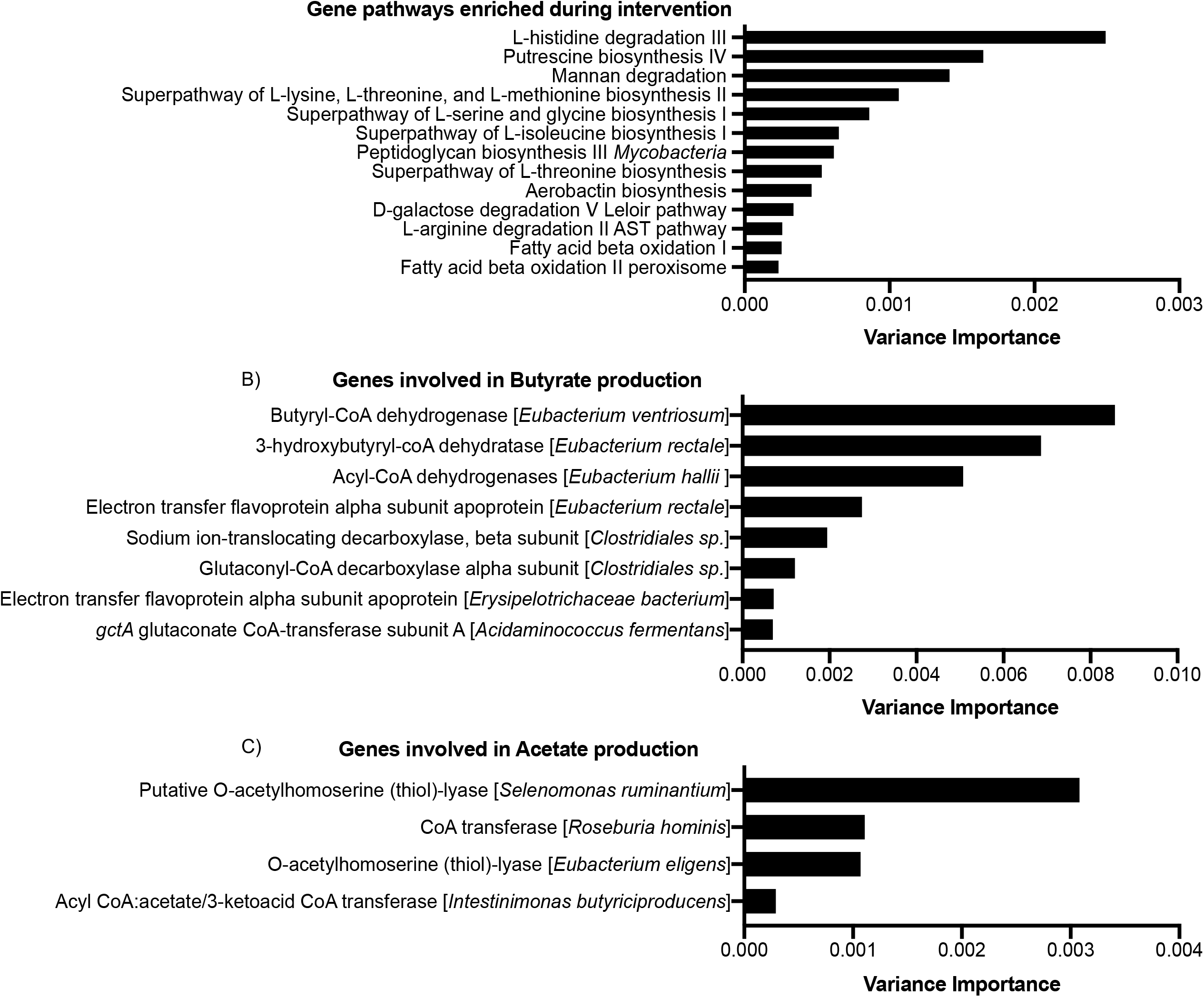
The IBD-AID increases the microbiome capacity for SCFA production. Bar plots represent the variance importance of the: A) gene pathways, B) genes involved in butyrate production, or C) genes involved in acetate production; that were enriched during the intervention (BH p-value > 0.05).

As previously done by us (32), we created specific databases that included all the bacteria genes involved in the production of the main 3 SCFAs in the gut: butyrate, acetate, and propionate. We found that subjects completing the intervention displayed an increased abundance of specific genes involved in the production of butyrate, mostly from members of the *Clostridia* class (**Figure 4B**); and acetate, specifically from *Roseburia hominis* and *Eubacterium eligens* species (**Figure 4C**). Lastly, genes linked to propionate production were not enriched during the intervention. Contrary, genes involved in propionate metabolism were augmented in baseline along with *Ruminococcus torques, Flavonifractor plautii*, and *Parabacteroides distasonis* (data not shown).

In sum, the diet-dependent changes of the microbiome were accompanied by increased microbial genomic capacity for butyrate and acetate metabolism.

### Foods responsible for the microbiome changes

To do this we first apply mixed effect random forest modeling to predict the abundance of each microbiome species as a function of the number of servings for each food category. To control for the effect of non-diet and other clinical covariates (i.e., age, gender, and BMI) we included them in the model as additional fixed effects. Similarly, to account for possible diagnosis-specific effects, we included in the model as additional fixed effects the interaction between every food category and the diagnosis. To determine the significance of the determined associations we run Permutated Importance (PIMP) analysis (see Methods). To determine the strength and direction of the association we then run Repeated Measure Correlations on the associations with a PIMP-associated p-value less than 0.05. We investigated the bacteria: food correlation of the top bacterial species enriched at either baseline or intervention in CD and UC participants, plus *B. dorei* and *P. distasonis* both enriched respectively at intervention or baseline regardless of disease phenotype (**Figure 5**). As expected, consumption of prebiotics, probiotics, and beneficial foods positively correlated with *Clostridia* and *Bacteroides* species enriched at intervention but negatively correlated with species enriched at baseline. Opposite correlations were observed with the consumption of adverse foods. A list with all the significant bacteria: food correlations are shown in Supplementary Table 6. Of interest, increased consumption of lean animal proteins (included in beneficial foods) has a negative correlation with *Roseburia hominis* in UC but not in CD participants.

**Figure 5.**
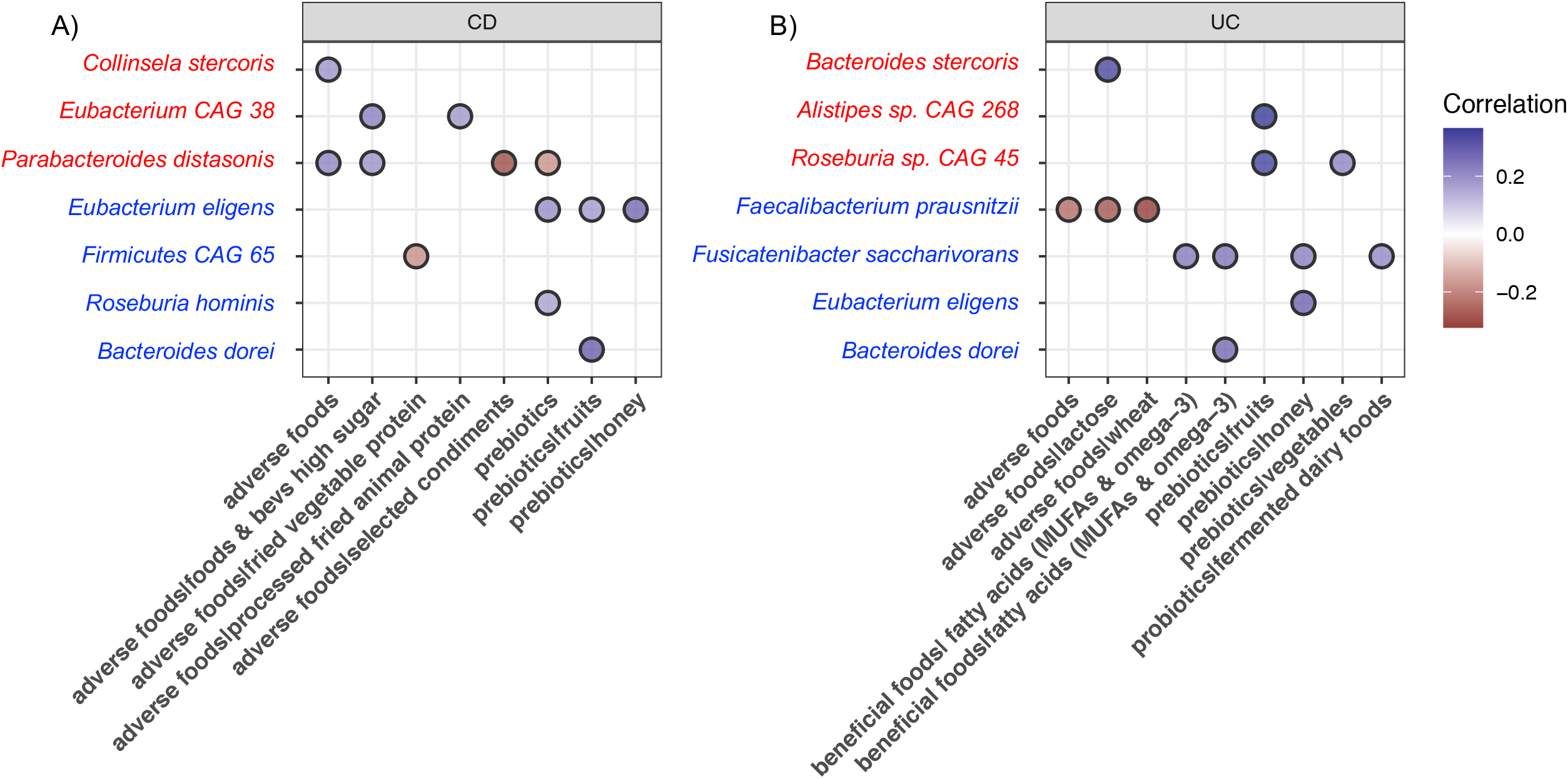
Significant correlations of foods with bacterial species enriched at baseline (red) or intervention (blue) in A) CD participants and B) UC participants.

Overall, our results show that increased consumption of prebiotics, probiotics, and beneficial foods do favor *Clostridia* and *Bacteroides* species depleted in IBD patients. We observed that the effect of some foods on bacteria abundance is dependent on disease phenotype.

### Immune modulation after the IBD-AID

We obtained blood samples from 9 patients before and after the intervention to measure circulating cytokines relevant to inflammation (**Supplementary Figure 3**). Out of 14 cytokines assessed, we found that the levels of the pro-inflammatory interleukin 8 (IL-8), tended to decrease following intervention (**Figure 6A**). Conversely, granulocyte macrophage-colony-stimulating factor (GM-CSF), also tended to increase after intervention (**Figure 6B**). Low levels of GM-CSF have been associated with IBD pathogenesis (33-39).

**Figure 6.**
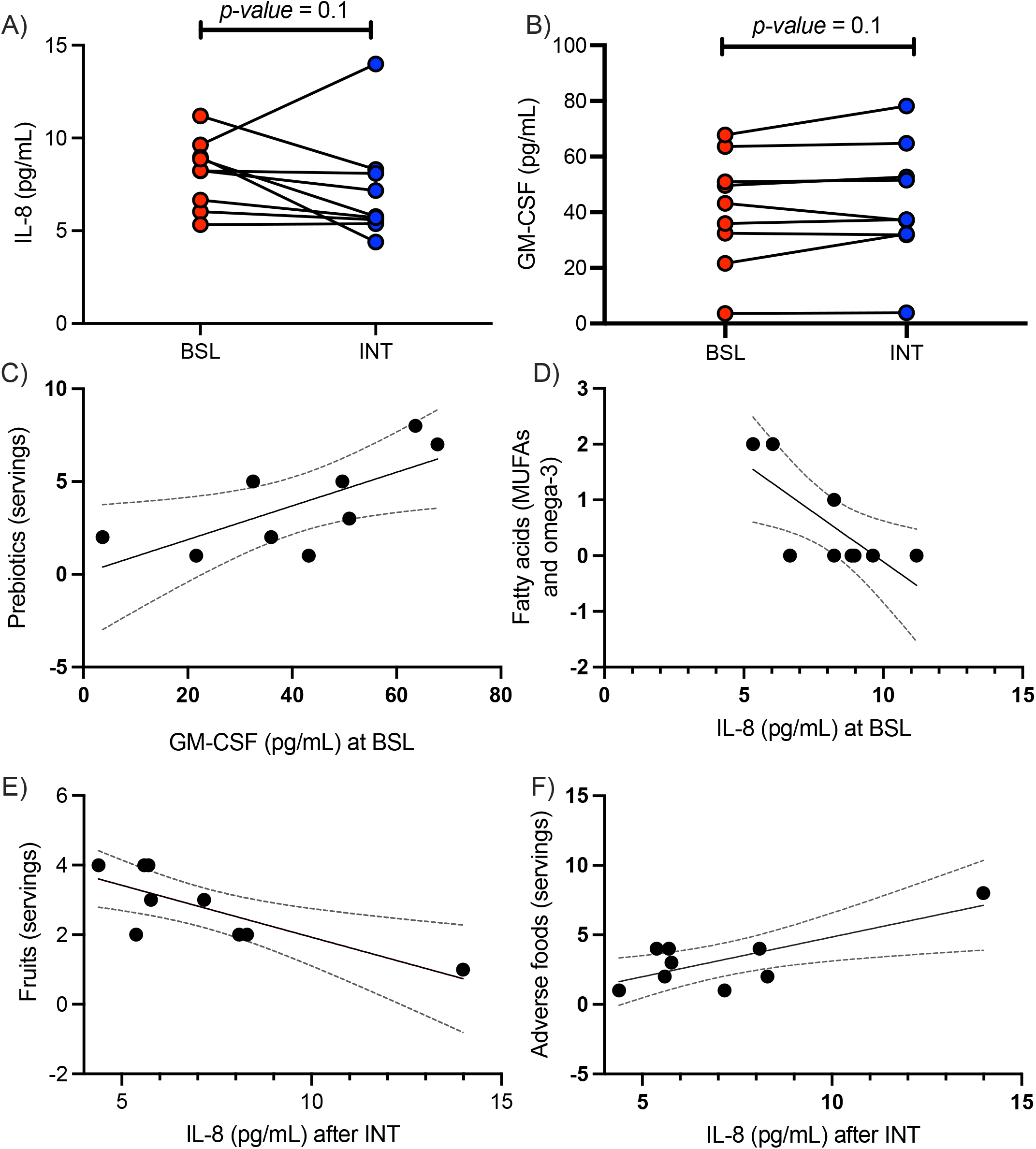
Levels of A) IL-8 significantly decrease from baseline (n = 9, red circles) to intervention (n = 9, blue circles) while B) GM-CSF significantly increases after intervention (Wilcoxon-matched pairs-signed rank test, p-value = 0.1). At baseline, increased consumption of C) prebiotics correlated with high levels of GM-CSF, and D) healthy fatty correlated with lower levels IL-8. At the intervention, increased consumption of E) fruits correlated with lower levels of IL-8, while F) adverse foods correlated with higher levels of IL-8 (Simple linear regression, *p-values* <0.05. Dotted lines represent 95% confidence intervals).

We then determine the correlation of the average of food intakes with the levels of IL-8 or GM-CSF at the baseline and intervention (**Figure 6C-F**). We observed that levels of GM-CSF at baseline positively correlated with increased consumption of prebiotics (Simple linear regression: slope= 0.09; 95% confidence intervals 0.01 to 0.17; *p value*: 0.03). Also at baseline, levels of IL-8 negatively correlated with consumption of fatty acids (Simple linear regression: slope= -0.35; 95% confidence intervals -0.64 to -0.06; *p value*: 0.02). After the intervention, consumption of fruits (Simple linear regression: slope= -0.29; 95% confidence intervals -0.51 to -0.09; *p value*: 0.01) and adverse foods (Simple linear regression: slope= 0.57; 95% confidence intervals 0.13 to 1.01; *p value*: 0.01) negatively and positively correlated with IL-8 levels, respectively.

In sum, participants on the IBD-AID exhibited a trend to lower levels of pro-inflammatory IL-8, which was correlated with decreased intake of adverse foods and increase consumption of fatty acids and fruits. Levels of colitis protective GM-CSF are increased after IBD-AID and it’s correlated with high consumption of prebiotics.

## DISCUSSION

Here we demonstrate that IBD patients can rapidly and dramatically change their diet and in doing so revert dysbiosis and modulate important cytokines driving IBD pathogenesis. Specifically, our results demonstrate that increased consumption of prebiotics (fiber-rich foods such as fruits, vegetables, oats, and honey), probiotics (fermented dairy products), and beneficial foods (lean animal protein and omega 3 fatty acids) can favor potent SCFA-producing *Clostridia* and *Bacteroides* species with known anti-inflammatory activity (9, 11, 21, 40-49) and which are known to be reduced in numerous cohorts of IBD patients across the world (5, 6, 10, 50-58).

High-fiber diets are related to healthy-like microbiomes (59-61) and have received increasing attention to reducing IBD risk and symptoms (18, 62-65). Here, increased consumption of fruits, vegetables, honey, and oats favored bacteria commonly depleted in IBD, namely: *Roseburia hominis, F. praustnizii, E. eligens, F. saccharivorans, B. dorei, B. vulgatus*. As with increased intakes of prebiotics vegetables and fruits, vegan and vegetarian diets have been also associated with increased microbiome capacity for biosynthesis of essential amino acids (66) that leads to the production of butyrate and acetate (67-72). Similarly, we observed that microbial gene pathways for biosynthesis of amino acids along with pathways involved in butyrate and acetate production were enriched along with participants’ increase in fruits and vegetable consumption.

Fermented foods have been shown to play an important role in microbiome diversity and concomitant immune tone on the host (73). In our cohort, there was a modest increase in the intake of fermented foods (average 0.5 servings per week), especially dairy products (i.e., yogurt, kefir). Despite the modest change in consumption, fermented dairy products in UC patients correlated with increased abundance of *Fusicatenibacter saccharivorans*, a bacteria known to be depleted on active UC patients (74).

Within the beneficial foods, we found that increased intakes of MUFAs and omega-3 fatty acids also support potent SCFA-producing *Clostridia* and *Bacteroides* species. Omega-3 fatty acids have previously been found to reduce dysbiosis (75-80). Of interest, we observed that lean animal proteins, included in the beneficial foods for the IBD-AID, are negatively associated with *Roseburia hominis* in UC patients. In line with this observation the International Organization for the Study of Inflammatory Bowel Diseases, recommends limiting animal protein intake for UC patients but not CD (81). Together, this highlights the importance of personalization of diet therapy based on the patient’s disease manifestation. Moreover, our results also emphasize the importance of a dietary approach for treating IBD that includes *adding* needed food components such as prebiotics, probiotics, and beneficial foods.

As expected, avoidance of foods also played an important role in shifting the microbiome during the intervention. We found that bacteria enriched at intervention (i.e., *Faecalibacterium prausnitzii, Firmicutes CAG 65*) negatively correlated with consumption of adverse foods. Conversely, bacteria species enriched at baseline (i.e, *Collinsela stercoris, Parabacteroides distonis*) were positively correlated with increased intakes of adverse foods. The abundance of *Collinsella* species has previously been associated with low-fiber diets (59, 82) and processed foods (83). Moreover, *Collinsella* sp. isolated from IBD patients conferred significant susceptibility to colitis in germ-free mice (84). Thus, we speculate that the increase of prebiotics rich in fiber and reduction of adverse processed foods at intervention reduces *Collinsella* fitness (83); leading to an outgrowth of SCFA-producing bacteria as a result of “new” nutrient availability (85-87). *Parabacteroides distonis*, enriched at baseline in both CD and UC participants in this cohort, have been also found abundant in IBD patients (88) and it has been implicated in worsening DSS-induced colitis in mice (89).

Along with changes in the microbiome, we also provide evidence of reduction of IL-8, a marker of inflammation, after intervention with IBD-AID. IL-8 is a potent neutrophil chemoattractant that is elevated in patients with either CD or UC and is correlated with mucosal inflammation (90-93). Reduction of IL-8 levels positively correlated with reduced consumption of adverse foods and increase intakes of fruits and healthy fatty acids (MUFAs and omega-3). On the other hand, subjects demonstrated increased levels of GM-CSF following the intervention. GM-CSF is a cytokine involved in myeloid cell development and maturation and dendritic cell differentiation. There is growing evidence that lower levels of GM-CSF are associated with the pathogenesis of CD (33-39). Here, we found that increased consumption of prebiotics was positively associated with levels of GM-CSF.

In conclusion, we demonstrate that the IBD-AID can favor bacteria commonly depleted in IBD patients and are key for maintaining immune tolerance and homeostasis in the gut via SCFA production. This short-term manipulation of the microbiome through diet resulted in modulation of the immune tone. Moreover, the results provide evidence for further adjustments of the foods allowed on the IBD-AID according to the disease phenotype.

## METHODS

### Power Calculation

The primary outcome is to evaluate the effect of the IBD-AID in the increased abundance of SCFA-producing bacteria. Using Monte-Carlo simulations of empirical power and type-I-error for a Wilcoxon-signed rank test (paired; R package MKpower (94)) we determined that 10 independent subjects (pre-post) will detect 0.005 +/-0.005 changes in the relative abundance of bacteria, with a power of 0.80. Our secondary outcome was associations between IBD-AID food categories and the microbiome. An unweighted Spearman correlation power analysis (R package genefu (95)) determined that 14 independent samples will be sufficient to achieve a significance of 0.05 and a correlation coefficient of 0.1.

### Participants

We recruited 25 subjects with an IBD diagnosis of either Crohn’s disease (CD) or ulcerative colitis (UC, **Figure 1A**). Of the 19 subjects who completed the study, 7 CD and 2 UC subjects were in remission at enrollment. The remaining 10 subjects exhibited either mild, moderate, or active disease. Exclusion criteria included: use the antibiotic within 3 months at the time of recruitment, presence of infection precipitating colitis (i.e., *C. difficile*), and pregnancy. For more inclusion and exclusion criteria see **Supplementary Table 1**. None of the participants reported antibiotic treatment during the study. The study was approved by the IRB at UMASS (Docket Number H00008033). ClinicalTrial.gov registry website: https://clinicaltrials.gov/ct2/show/NCT04757181 Trial number: NCT04757181.

### The IBD-AID

As published elsewhere (19), the IBD-AID supports the avoidance of certain carbohydrates (sucrose and starches) from the original SCD (96). Before the mSCD, the IBD-AID was the first IBD diet to include oats as a source of fiber. The IBD-AID encourage the increased intakes of monounsaturated and polyunsaturated omega-3 and fatty acids while decreasing other saturated fats and eliminating trans-fatty acids (62, 75, 97-103). Moreover, the IBD-AID eliminates the consumption of processed and ultra-processed foods which have been associated with IBD risk (104). The IBD-AID includes prebiotics: foods rich in non-digestible fiber that serve as food for beneficial bacteria colonizing the colon (105, 106). Epidemiological evidence (107-110) and results from a recent clinical trial study (73) support the role of fermented foods containing live active bacteria (probiotics) in health, microbiome diversity, and an anti-inflammatory immune status (73). Thus, the IBD-AID also encourages the consumption of probiotics. Finally, to avoid nutrient deficiencies that could be caused by restrictive, the IBD-AID also encourages the intake of nutritious foods recommended by the Dietary Guidelines for Americans (111); which includes a variety of foods rich in essential vitamins and minerals. The diet can be prepared at home and is designed to be healthful long-term for the entire family.

### Intervention

We conducted a prospective, single-arm, pre-post intervention trial. After a baseline period of 6 weeks, the dietary intervention was initiated and continued for 8 weeks (**Figure 1B)**. To receive dietary instructions, subjects met in person with trained nutritionists at the beginning of the intervention and completed at least one counseling session per week throughout the 8-weeks intervention period.

### Dietary Assessment

To measure food intake we developed: ‘24-hour IBD-AID Food Query’, which was programmed in REDCap and consists of 240 food items grouped in four main food categories: prebiotic foods (fruits, vegetables, legumes, oats, and honey); probiotic foods (fermented dairy products, and fermented non-dairy foods); beneficial foods (fatty acids rich in monounsaturated and omega-3 polyunsaturated fatty acids, vegetable and lean animal proteins); and adverse foods (wheat, corn, lactose, high fat animal and vegetable proteins, processed fried foods, artificial sweeteners, foods, and beverages high in sugar, high-fat processed foods, selected starchy vegetables, selected gluten free grains, and certain condiments). Alcohol consumption was accounted for in a separate category. A link to the electronic 24-hour IBD-AID Food Query was sent to the participants to be filled out 3 times per week. The serving sizes recorded on each 24-hour IBD-AID Food Query were assigned to the individual food categories mentioned above and the serving sizes reported were averaged per week for analysis.

### Sample collection

Subjects were provided materials and instructions for at-home self-collection using OMNIgene•GUT collection kits (#OM-200, DNA Genotek Inc., Ottawa, Canada). We also obtained blood samples at baseline and the end of the intervention. Once in the laboratory, samples were aliquoted and then stored at -80°C until processed.

### DNA isolation and sequencing

DNA isolation was performed using the MagAttract PowerSoil DNA Kit (#27100-4-EP, Qiagen, Germantown, MD, USA) on Eppendorf epMotion 5075 liquid handlers following the manufacturer’s instructions. Libraries for DNA sequencing were prepared using the Nextera XT DNA Library Preparation Kit (#FC-131-1096, Illumina, San Diego, CA, USA) and were sequenced on the Illumina NextSeq 500 platform using 150-nt paired-end reads. We obtained an average of 4,926,661 reads per sample. Read data were quality trimmed and filtered of host DNA using KneadData (version 0.7.2; https://bitbucket.org/biobakery/kneaddata/wiki/Home) against a prebuilt bowtie2 index for the human genome, hg19. All the filtered sequences generated were deposited in NCBI, BioProject: PRJNA642308.

### Metagenomic profiling

We performed shotgun metagenomic sequencing of stool samples as previously described by us and others (32, 112-114). Community composition was profiled using MetaPhlan2 (version 2.9.14; database mpa_v292_CHOCOPhlAn_201901) (115). To assess the abundance of microbiota-encoded metabolic pathways we used HUMAnN2 (version 2.8) (116). We used ShortBRED (117) to profile metagenomics reads for the abundance of proteins involved in the production of SCFAs (e.g., butyrate, acetate, propionate) as we have previously described (32, 118).

### Inflammatory markers

We used the Discovery Assay^®^ Human High Sensitivity T-Cell Discovery Array 14-Plex (#HDHSTC14, Eve Technologies Corp, Calgary, Canada) to simultaneously quantified 14 cytokine/chemokine/growth involved in inflammation.

### Mathematical modeling

#### Microbiome associations with study periods

To determine the bacterial species impacted by the IBD-AID we applied mixed-effect random forest classification by adapting the MERF R engine (119). This framework enables to account for the repeated sampling nature of the dataset and is appropriately suited for this type of “large p, small n” multi-omics dataset common in clinical research (120). We classify a sample *i* from patient *s* as Intervention *vs*. Baseline (Y_is_ = 1,0) as a function of microbiome abundance in that sample as a fixed effect (X_**is**_) and controlling for the individual patient as a random effect (Z): *Y*_*ij*_ = *f*(*X*_*i*_) + *b*_*j*_*Z* + *ϵ*. Compared to traditional linear mixed-effect modeling regression here *f* is a general function that is learned using a random forest model. The expectation-maximization algorithm runs via alternative optimization, in which, at the turn, one parameter is fitted while the other ones are fixed with the process running until convergence (119). This analysis was repeated using as predictors species abundances, metabolic pathways abundances, and SCFAs pathways, independently. Permutated importance (PIMP) analysis was used to estimate the significance of each microbiome feature in the classification analyses (112, 121).

#### Microbiome associations with food categories

We determined the effect of food categories on the microbiome by first applying mixed-effect random forest regression modeling while also controlling for other clinical and not-diet related covariates (i.e., age, gender, and BMI) (119). To account for diagnosis (UC, CD) -dependent effects of food categories on the microbiome, we also consider the interactions between food-category (as number of servings, numerical) and the diagnosis (categorical) in the modeling. As above, PIMP analysis was used to estimate the significance of each model covariate in predicting the abundance of every modeled microbial feature (121),(112). For the food covariates displaying a PIMP-associated p-value < 0.05, we run repeated measure correlation for UC and CD individuals independently to determine the direction and significance of the identified association.

### Statistical analysis

We used Prism 9 to perform the statistical analyses. We used the Mann-Whitney test with individual ranks computed per comparison of food intakes by study phase using the two-stage linear step-up procedure of Benjamini, Krieger, and Yekutieli correction. Wilcoxon matched-pairs signed-rank was used to evaluate differences in cytokine concentration in serum before and after the diet intervention; due to the low sample size (n=9), *p values* of 0.1 are reported as trends. Simple linear regressions were calculated between the average of intakes of each food category and the levels of cytokines at each study period. We used the R package Phyloseq v1.19.1 (122) to calculate the Shannon diversity index (123, 124) and Bray-Curtis dissimilarity. Statistical significance of Bray-Curtis distances was assessed using PERMANOVA in R (125).

## Data Availability

All data produced in the present study are available upon reasonable request to the authors.

## Abbreviations used in this paper

(IBD): Inflammatory bowel disease
(CD): Crohn’s disease
(UC): Ulcerative colitis
(IBD-AID): Inflammatory bowel disease Anti-Inflammatory Diet
(SCFAs): Short-chain fatty acids
(MD): Mediterranean diet
(SCD): Specific Carbohydrate Diet

## AUTHORS CONTRIBUTIONS

BO, EO, DW, RP, BMC, and AMC – study conceptualization and methodology. BO, CC, RM, MMM, CM, DC, RP – investigation and data curation. BO, VB, EO, DW, DC, CF, SB, and AMC – formal analysis of data. BO, VB, DW, BMC, and AMC – writing – original draft, review & editing. AMC – funding acquisition. BO and AMC –supervision.

## ACKNOWLEDGMENTS

We are grateful for the effort and willingness all the study participants demonstrated throughout the study. We thank Kanishka Bhattacharya for patient referral to the study. We appreciate Yurima Guillarte-Walker’s help in identifying patients for the study. The study was funded by the American Gastroenterological Association and partially by the Leona M. and Harry B. Helmsley Charitable Trust.

## Notes

**Authors Conflict of Interest** Beth A. McCormick (BAM) is a coinventor on a patent application (PGT/US 18/42116). She, along with her academic institution, stands to gain financially through potential commercialization outcomes resulting from activities associated with the licensing of that intellectual property. The remaining authors do not have any conflict of interest to disclose.

### Competing Interest Statement

Beth A. McCormick (BAM) is a coinventor on a patent application (PGT/US 18/42116). She, along with her academic institution, stands to gain financially through potential commercialization outcomes resulting from activities associated with the licensing of that intellectual property. The remaining authors do not have any conflict of interest to disclose.

### Clinical Trial

NCT04757181

### Funding Statement

This study was funded by the American Gastroenterological Association: Research Scholar Award, the University of Massachusetts Medical School: Faculty Diversity Scholar, and partially by the Leona M. and Harry B. Helmsley Charitable Trust.

### Author Declarations

The study was approved by the IRB at UMASS (Docket Number H00008033). Signed informed consent was obtained from all study participants

