## Supplementary Figure Legends for "DIETARY MANIPULATION OF THE GUT MICROBIOME IN INFLAMMATORY BOWEL DISEASE PATIENTS: PROOF OF CONCEPT"

**Supplementary Figure 1.** A) Shannon diversity and B) T-distributed stochastic neighbor embedding analysis shows that both CD and UC participants showed a personalized microbiome that clustered individually rather than by disease phenotype.

**Supplementary Figure 2**. A) Shannon diversity and B) tSNE clustering for samples at Baseline (BSL) and during Intervention (INT)

**Supplementary Figure 3**. Levels of circulatory inflammatory markers at baseline (BSL; N=9, red circles) and at the end of the intervention (INT; N=9, blue circles).
