## Supplementary figures and images for "DIETARY MANIPULATION OF THE GUT MICROBIOME IN INFLAMMATORY BOWEL DISEASE PATIENTS: PROOF OF CONCEPT"

### Supplementary Figure 1

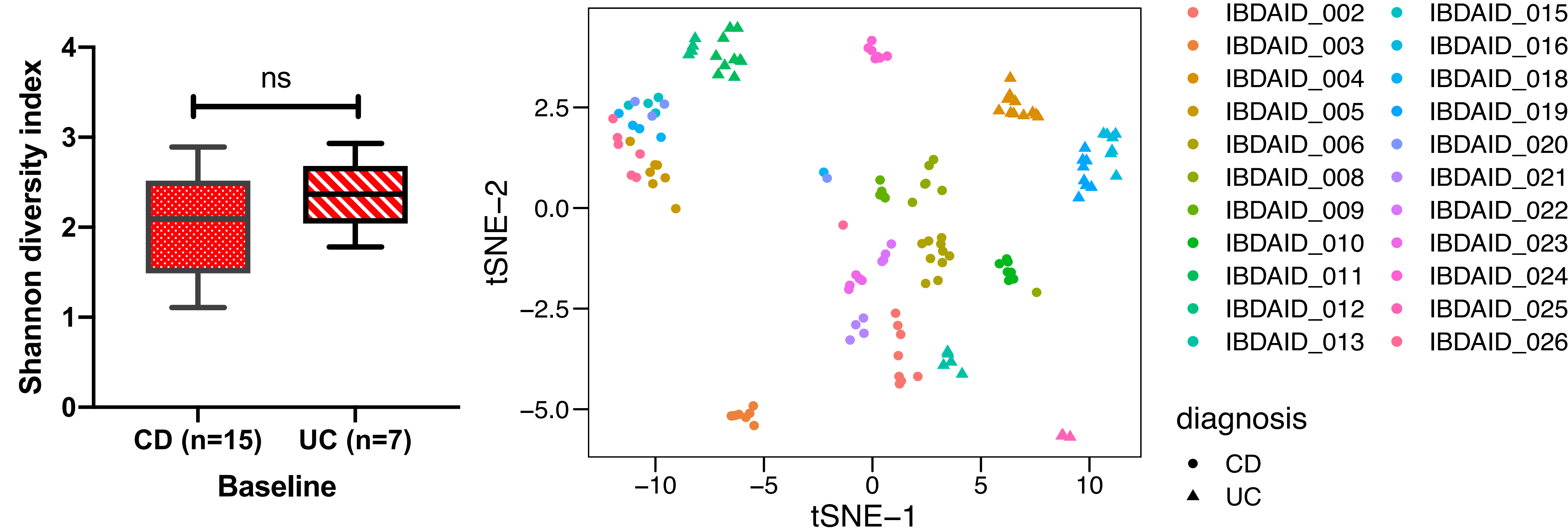

### Supplementary Figure 2

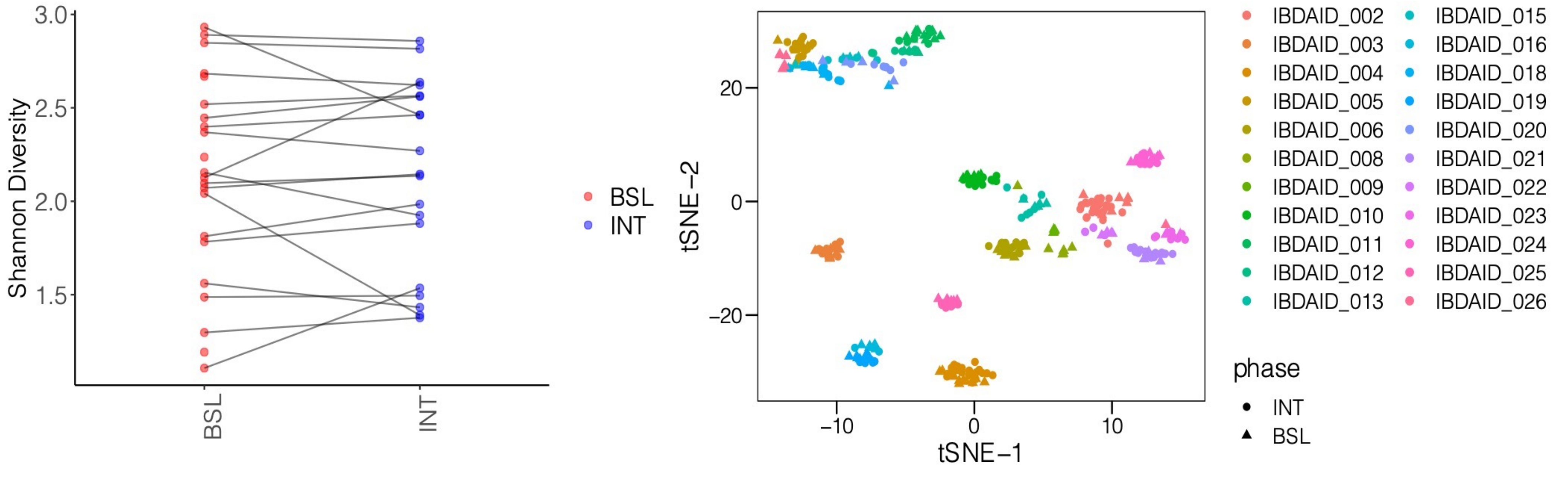

### Supplementary Figure 3

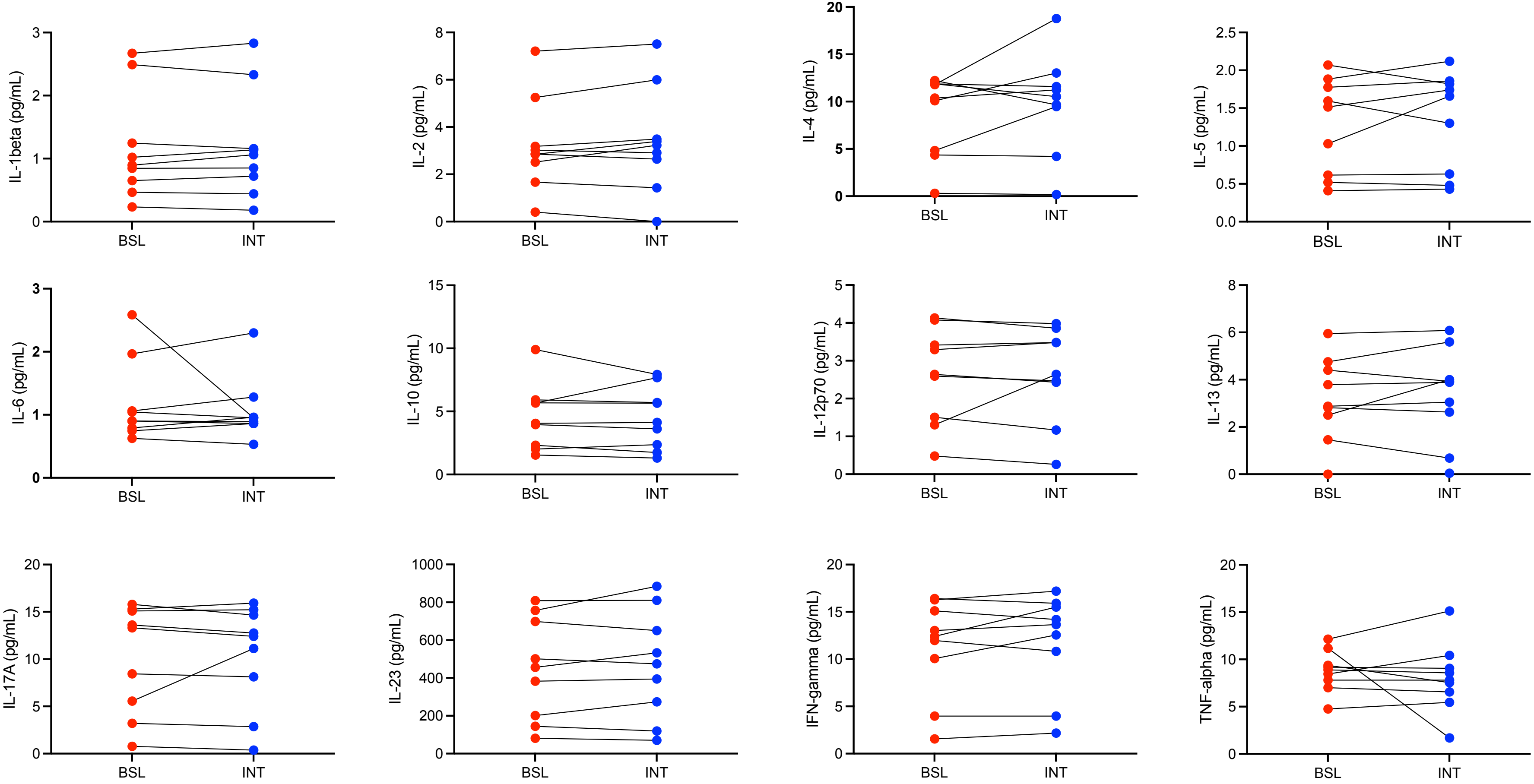
