## Supplementary Tables for "DIETARY MANIPULATION OF THE GUT MICROBIOME IN INFLAMMATORY BOWEL DISEASE PATIENTS: PROOF OF CONCEPT"

**Supplementary Table 1**. Inclusion and exclusion criteria for patient enrollment for the IBD-AID™ intervention between 2017 and 2018

Inclusion criteria:

- age 15-70 years old
- Willingness and capacity to significantly change diet
- Willing and able to comply with scheduled in-person or over the phone study activities
- Primary care provider permission for IBD patients to participate in the intervention is required for participation in the study
- Evidence of personally signed and dated informed consent document indicating that the subject has been informed of all pertinent aspects of the study
- For children aged 15 and above, the assent of the child and consent of parents or legal guardians will be required.
- Diagnosis of IBD: Crohn's disease or ulcerative colitis

Exclusion criteria:

- Use of antibiotics within 3 months at the time of consent
- Presence of infection precipitating the colitis
- Medically unable to give consent
- Prisoners
- Subjects who do not speak English.
- Subjects who self-report current pregnancy
- On heparin or Coumadin
- Hct < 24

**Supplementary Table 2**. Mean food intakes reported on the 24-hour IBD-AID Food Query during baseline by CD and UC participants (Mann-Whitney test, Bonferroni-Dunn correction for multiple comparisons).

| | Mean intakes at BSL (CD=14) | Mean intakes at BSL (UC=7) | Difference | Adjusted P value |
|---|---|---|---|---|
| ***Prebiotics*** | 3.755 | 4.556 | -0.8008 | >0.999999 |
| Fruits | 1.132 | 2.028 | -0.8957 | >0.999999 |
| Vegetables | 2.283 | 2.278 | 0.005241 | >0.999999 |
| Oats | 0.2642 | 0.2222 | 0.04193 | >0.999999 |
| Honey | 0.1887 | 0 | 0.1887 | 0.250726 |
| ***Probiotics*** | 1.075 | 1.111 | -0.03564 | >0.999999 |
| Fermented foods (no dairy) | 0.1698 | 0.1389 | 0.03092 | >0.999999 |
| Fermented dairy foods | 0.8679 | 1 | -0.1321 | >0.999999 |
| ***Beneficial foods*** | 3.283 | 4.75 | -1.467 | **0.02559** |
| Fatty acids (MUFAs & Omega-3) | 0.7547 | 1.528 | -0.7731 | 0.255502 |
| Vegetable protein | 0.6038 | 0.3333 | 0.2704 | >0.999999 |
| Animal protein (lean) | 1.811 | 2.611 | -0.7998 | 0.094033 |
| ***Adverse foods*** | 11.98 | 13.61 | -1.63 | >0.999999 |
| Wheat | 2.642 | 2.389 | 0.2526 | >0.999999 |
| Lactose | 2.057 | 1.917 | 0.1399 | >0.999999 |
| High fat animal protein | 0.1509 | 0.3333 | -0.1824 | >0.999999 |
| Processed fried animal protein | 0.5849 | 0.5833 | 0.001572 | >0.999999 |
| Fried vegetable protein | 0.5472 | 1 | -0.4528 | >0.999999 |
| Artificial sweeteners | 1.094 | 0 | 1.094 | **0.001775** |
| Food beverages high sugar | 2.528 | 4.472 | -1.944 | >0.999999 |
| High fat processed foods | 1.34 | 1.111 | 0.2285 | >0.999999 |
| Selected gluten free grains | 0.7358 | 0.75 | -0.01415 | >0.999999 |
| Corn and starchy vegetables | 0.3019 | 0.1944 | 0.1074 | >0.999999 |
| Selected condiments | 0.2075 | 1.056 | -0.848 | 0.083299 |
| ***Alcohol*** | 0.4717 | 0.4167 | 0.05503 | >0.999999 |

**Supplementary Table 3**. Mean food intakes reported on the 24-hour IBD-AID Food Query by CD participants during the two periods of the study: baseline and intervention (Mann-Whitney test, Bonferroni-Dunn correction for multiple comparisons. **In bold**: significant Adjusted p values).

| | Mean intakes at BSL (CD, n=14) | Mean intakes at INT (CD, n=12) | Difference | Adjusted P value |
|---|---|---|---|---|
| ***Prebiotics*** | 3.755 | 7.417 | 3.662 | **<0.000001** |
| Fruits | 1.132 | 2.292 | 1.160 | **0.000029** |
| Vegetables | 2.283 | 3.667 | 1.384 | **0.002472** |
| Oats | 0.2642 | 0.7917 | 0.5275 | **0.000379** |
| Honey | 0.1887 | 0.7361 | 0.5474 | **0.002382** |
| ***Probiotics*** | 1.075 | 1.444 | 0.3690 | 0.118669 |
| Fermented foods (no dairy) | 0.1698 | 0.2361 | 0.06630 | >0.999999 |
| Fermented dairy foods | 0.8679 | 1.250 | 0.3821 | **0.026864** |
| ***Beneficial foods*** | 3.283 | 5.417 | 2.134 | **<0.000001** |
| Fatty acids (MUFAs & Omega-3) | 0.7547 | 2.514 | 1.759 | **<0.000001** |
| Vegetable protein | 0.6038 | 0.5694 | -0.03433 | >0.999999 |
| Animal protein (lean) | 1.811 | 2.319 | 0.5081 | 0.099246 |
| ***Adverse foods*** | 11.98 | 3.569 | -8.412 | **<0.000001** |
| Wheat | 2.642 | 0.3889 | -2.253 | **<0.000001** |
| Lactose | 2.057 | 0.2778 | -1.779 | **<0.000001** |
| High fat animal protein | 0.1509 | 0.09722 | -0.05372 | >0.999999 |
| Processed fried animal protein | 0.5849 | 0.2083 | -0.3766 | **0.006396** |
| Fried vegetable protein | 0.5472 | 0.1389 | -0.4083 | **0.000161** |
| Artificial sweeteners | 1.094 | 1.042 | -0.05267 | >0.999999 |
| Food beverages high sugar | 2.528 | 0.4583 | -2.070 | **<0.000001** |
| High fat processed foods | 1.340 | 0.3333 | -1.006 | **<0.000001** |
| Selected gluten free grains | 0.7358 | 0.4167 | -0.3192 | 0.278332 |
| Corn and starchy vegetables | 0.3019 | 0.08333 | -0.2186 | **0.023542** |
| Selected condiments | 0.2075 | 0.1528 | -0.05477 | >0.999999 |
| ***Alcohol*** | 0.4717 | 0.6667 | -0.1950 | 0.206838 |

**Supplementary Table 4**. Mean food intakes reported on the 24-hour IBD-AID Food Query by UC participants during the two periods of the study: baseline and intervention (Mann-Whitney test, Bonferroni-Dunn correction for multiple comparisons. **In bold**: significant Adjusted p values).

| | Mean intakes at BSL (UC, n=7) | Mean intakes at INT (UC, n=7) | Difference | Adjusted P value |
|---|---|---|---|---|
| ***Prebiotics*** | 4.556 | 7.683 | 3.127 | **0.000540** |
| Fruits | 2.028 | 3.122 | 1.094 | **0.000737** |
| Vegetables | 2.278 | 3.366 | 1.088 | **0.031425** |
| Oats | 0.2222 | 0.6585 | 0.4363 | 0.183204 |
| Honey | 0.000 | 0.4634 | 0.4634 | **0.000108** |
| ***Probiotics*** | 1.111 | 1.854 | 0.7425 | **0.028744** |
| Fermented foods (no dairy) | 0.1389 | 0.3659 | 0.2270 | 0.398627 |
| Fermented dairy foods | 1.000 | 1.488 | 0.4878 | 0.363855 |
| ***Beneficial foods*** | 4.750 | 7.390 | 2.640 | **0.002779** |
| Fatty acids (MUFAs Omega-3) | 1.528 | 3.610 | 2.082 | **0.000683** |
| Vegetable protein | 0.3333 | 0.9512 | 0.6179 | **0.020355** |
| Animal protein (lean) | 2.611 | 2.634 | 0.02304 | >0.999999 |
| ***Adverse foods*** | 13.61 | 3.146 | -10.46 | **0.000077** |
| Wheat | 2.389 | 0.1951 | -2.194 | **0.000002** |
| Lactose | 1.917 | 0.4146 | -1.502 | **0.000501** |
| High fat animal protein | 0.3333 | 0.09756 | -0.2358 | 0.116233 |
| Processed fried animal protein | 0.5833 | 0.3171 | -0.2663 | 0.572163 |
| Fried vegetable protein | 1.000 | 0.09756 | -0.9024 | **0.000035** |
| Artificial sweeteners | 0.000 | 0.000 | 0.000 | >0.999999 |
| Food beverages high sugar | 4.472 | 1.024 | -3.448 | **0.000135** |
| High fat processed foods | 1.111 | 0.3415 | -0.7696 | **0.017319** |
| Selected gluten free grains | 0.7500 | 0.4390 | -0.3110 | >0.999999 |
| Corn and starchy vegetables | 0.1944 | 0.07317 | -0.1213 | >0.999999 |
| Selected condiments | 1.056 | 0.1463 | -0.9092 | **0.013611** |
| ***Alcohol*** | 0.4167 | 0.1951 | -0.2215 | 0.582709 |

**Supplementary Table 5**. Bacteria species enriched in each study period when comparing all, CD, or UC subjects. Mixed effect random forest classification analysis identified microbes that are affected by the intervention.

| Predictors | Aleatory Effect | Variable Importance | *p-value* | Favored at |
|---|---|---|---|---|
| ***Comparing all the subjects by study period*** | | | | |
| *Eubacterium sp. CAG: 38* | -0.020221592 | 0.009647596 | 0 | Baseline |
| *Collinsella stercoris* | -0.002637033 | 0.006988067 | 0 | Baseline |
| *Alistipes sp. CAG: 268* | -0.005031791 | 0.005206963 | 0 | Baseline |
| *Parabacteroides distasonis* | -0.004817476 | 0.005178225 | 0 | Baseline |
| *Veillonella parvula* | -0.021928166 | 0.004785468 | 0.01 | Baseline |
| *Collinsella intestinalis* | -0.002305167 | 0.004601943 | 0 | Baseline |
| *Bacteroides stercoris* | -0.004707766 | 0.004376012 | 0 | Baseline |
| *Bacteroides xylanisolvens* | -0.001843794 | 0.002585475 | 0 | Baseline |
| *Flavonifractor plautii* | -0.000396738 | 0.002105678 | 0.03 | Baseline |
| *Collinsella aerofaciens* | -0.001466835 | 0.002082137 | 0.02 | Baseline |
| *Anaeromassilibacillus sp. An250* | -0.007277254 | 0.002015971 | 0 | Baseline |
| *Blautia sp. CAG: 52* | -0.006407906 | 0.001671531 | 0 | Baseline |
| *Prevotella sp. CAG: 386* | -0.017450065 | 0.001516188 | 0.01 | Baseline |
| *Blautia wexlerae* | -0.001849405 | 0.001450893 | 0 | Baseline |
| *Eubacterium ramulus* | -0.00429964 | 0.001342406 | 0 | Baseline |
| *Fimbriiglobus ruber* | -0.008370366 | 0.001239691 | 0.01 | Baseline |
| *Bifidobacterium adolescentis* | -0.001536492 | 0.001010081 | 0.03 | Baseline |
| *Ruminococcus torques* | -0.000507828 | 0.000997965 | 0.01 | Baseline |
| *Bifidobacterium longum* | -0.002061039 | 0.000995134 | 0.03 | Baseline |
| *Parasutterella excrementihominis CAG: 233* | -0.001165813 | 0.000949952 | 0.01 | Baseline |
| *Alistipes finegoldii* | -0.003280036 | 0.000941651 | 0.03 | Baseline |
| *Olsenella scatoligenes* | -0.000612061 | 0.000882518 | 0.04 | Baseline |
| *Ruminococcus sp. CAG: 177* | -0.007241713 | 0.00081154 | 0.02 | Baseline |
| *Erysipelatoclostridium ramosum* | -0.003036319 | 0.000704616 | 0.02 | Baseline |
| *Firmicutes bacterium CAG: 83* | -0.000680442 | 0.000676266 | 0.04 | Baseline |
| *Bacteroides massiliensis* | -0.008932686 | 0.000630229 | 0.02 | Baseline |
| *Burkholderiales bacterium 1 1 47* | -0.000856456 | 0.00062904 | 0.04 | Baseline |
| *Bacteroides eggerthii* | -0.001078363 | 0.000602162 | 0.03 | Baseline |
| *Ruminococcus lactaris* | -0.000312182 | 0.000575367 | 0.03 | Baseline |
| *Tyzzerella sp.* | -0.002470637 | 0.000544081 | 0.02 | Baseline |
| *Lactobacillus rogosae* | -0.001441806 | 0.00038365 | 0.04 | Baseline |

| | | | | |
|---|---|---|---|---|
| *Roseburia hominis* | 0.012338108 | 0.046508562 | 0 | Intervention |
| *Firmicutes bacterium CAG: 65* | 0.00315891 | 0.006661845 | 0 | Intervention |
| *Alistipes shahii* | 0.003267289 | 0.005492278 | 0 | Intervention |
| *Eubacterium eligens* | 0.003353466 | 0.005368829 | 0 | Intervention |
| *Faecalibacterium prausnitzii* | 0.002566376 | 0.005122354 | 0.01 | Intervention |
| *Bacteroides vulgatus* | 0.001219837 | 0.003388989 | 0.03 | Intervention |
| *Blautia obeum* | 0.002407928 | 0.003189867 | 0 | Intervention |
| *Bacteroides dorei* | 0.000887902 | 0.002457491 | 0.01 | Intervention |
| *Bacteroides dorei CAG: 222* | 0.012119434 | 0.001627256 | 0 | Intervention |
| *Bilophila sp. 4 1 30* | 0.000831829 | 0.001269553 | 0.03 | Intervention |
| *Alistipes sp. HGB5* | 8.87E-05 | 0.001239861 | 0.04 | Intervention |
| *Bacteroides cellulosilyticus* | 0.004787761 | 0.001033409 | 0 | Intervention |
| *Clostridium sp. CAG: 62* | 0.00308838 | 0.00086535 | 0.01 | Intervention |
| *Eubacterium siraeum* | 0.006964921 | 0.000833574 | 0.01 | Intervention |
| *Enterorhabdus caecimuris* | 0.002049022 | 0.000788621 | 0.04 | Intervention |
| *Intestinimonas butyriciproducens* | 0.00071432 | 0.000740984 | 0.04 | Intervention |
| *Clostridium sp. CAG: 217* | 0.000920946 | 0.000572536 | 0.03 | Intervention |
| *Clostridium sp. CAG: 75* | 0.001030448 | 0.000563212 | 0.03 | Intervention |
| ***Comparing CD subjects by study period*** | | | | |
| *Collinsella stercoris* | -0.003074051 | 0.010260579 | 0 | Baseline |
| *Veillonella parvula* | -0.030412584 | 0.008238149 | 0 | Baseline |
| *Eubacterium sp. CAG: 38* | -0.020365619 | 0.006628911 | 0.01 | Baseline |
| *Parabacteroides distasonis* | -0.004786014 | 0.005675701 | 0.02 | Baseline |
| *Collinsella intestinalis* | -0.001851809 | 0.003566 | 0 | Baseline |
| *Collinsella aerofaciens* | -0.001472591 | 0.003044205 | 0.03 | Baseline |
| *Anaeromassilibacillus sp. An250* | -0.023986483 | 0.002530221 | 0 | Baseline |
| *Klebsiella pneumoniae* | -0.012451169 | 0.001993173 | 0.02 | Baseline |
| *Ruminococcus torques* | -0.000973994 | 0.001147678 | 0.03 | Baseline |
| *Blautia sp. CAG: 52* | -0.005344338 | 0.001129014 | 0.01 | Baseline |
| *Firmicutes bacterium CAG: 110* | -0.012253077 | 0.000900977 | 0 | Baseline |
| *Eubacterium ramulus* | -0.001854194 | 0.000378044 | 0.03 | Baseline |
| *Ruminococcus sp. CAG: 177* | -0.005096369 | 0.000366291 | 0.02 | Baseline |
| *Roseburia hominis* | 0.01563794 | 0.071176082 | 0 | Intervention |
| *Firmicutes bacterium CAG: 65* | 0.003201664 | 0.005020039 | 0 | Intervention |
| *Alistipes shahii* | 0.002078896 | 0.003630586 | 0.04 | Intervention |
| *Blautia obeum* | 0.002108586 | 0.003045422 | 0.02 | Intervention |
| *Bacteroides dorei CAG: 222* | 0.011267863 | 0.002683778 | 0 | Intervention |
| *Enterorhabdus caecimuris* | 0.004559663 | 0.00154646 | 0.03 | Intervention |
| *Eubacterium eligens* | 0.002073458 | 0.00152726 | 0.02 | Intervention |
| *Roseburia inulinivorans* | 0.001853925 | 0.001361285 | 0.04 | Intervention |

| | | | | |
|---|---|---|---|---|
| *Clostridium clostridioforme* | 0.00222368 | 0.001316033 | 0.03 | Intervention |
| *Erysipelatoclostridium ramosum* | 0.010150908 | 0.001034105 | 0.01 | Intervention |
| *Parabacteroides goldsteinii* | 0.00068132 | 0.000997028 | 0.03 | Intervention |
| *Clostridium sp. CAG: 75* | 0.002451757 | 0.000975271 | 0.04 | Intervention |
| *Bacteroides cellulosilyticus* | 0.002213315 | 0.000590466 | 0.01 | Intervention |
| *Lactobacillus rogosae* | 0.003044635 | 0.000533184 | 0.04 | Intervention |
| ***Comparing UC subjects by study period*** | | | | |
| *Bacteroides stercoris* | -0.0323232 | 0.03638115 | 0 | Baseline |
| *Alistipes sp. CAG: 268* | -0.0115237 | 0.01383164 | 0 | Baseline |
| *Roseburia sp. CAG: 45* | -0.0177914 | 0.01105742 | 0 | Baseline |
| *Prevotella sp. CAG: 386* | -0.0149628 | 0.00505348 | 0.03 | Baseline |
| *Parabacteroides merdae* | -0.0021774 | 0.00422862 | 0 | Baseline |
| *Ruminococcus gnavus* | -0.0052976 | 0.00317068 | 0.03 | Baseline |
| *Bacteroides xylanisolvens* | -0.0022562 | 0.00316538 | 0.03 | Baseline |
| *Parabacteroides distasonis* | -0.0024271 | 0.0031275 | 0.01 | Baseline |
| *Blautia wexlerae* | -0.0019697 | 0.00296462 | 0.04 | Baseline |
| *Firmicutes bacterium CAG: 65* | -0.0008306 | 0.00227063 | 0.04 | Baseline |
| *Bacteroides sp. CAG: 144* | -0.0081524 | 0.00204123 | 0.01 | Baseline |
| *Burkholderiales bacterium 1 1 47* | -0.0014054 | 0.00168602 | 0.04 | Baseline |
| *Eubacterium ramulus* | -0.0052917 | 0.00147794 | 0.04 | Baseline |
| *Clostridium aldenense* | -0.0098669 | 0.00113502 | 0.03 | Baseline |
| *Barnesiella intestinihominis* | -0.0036001 | 0.00100616 | 0.01 | Baseline |
| *Sutterella parvirubra* | -0.0009399 | 0.00088819 | 0.02 | Baseline |
| *Eubacterium hallii* | -0.0033539 | 0.00061488 | 0.03 | Baseline |
| *Ruminococcus sp. CAG: 177* | -0.0048299 | 0.00046903 | 0.02 | Baseline |
| *Eubacterium ventriosum* | -0.0001403 | 0 | 0 | Baseline |
| *Faecalibacterium prausnitzii* | 0.00556749 | 0.01340132 | 0 | Intervention |
| *Eubacterium eligens* | 0.00371664 | 0.0113536 | 0 | Intervention |
| *Fusicatenibacter saccharivorans* | 0.00276492 | 0.00887255 | 0 | Intervention |
| *Coprococcus comes* | 0.00425299 | 0.00452068 | 0.01 | Intervention |
| *Bacteroides dorei* | 0.0042783 | 0.00429151 | 0.02 | Intervention |
| *Bacteroides vulgatus* | 0.00102551 | 0.00296162 | 0.01 | Intervention |
| *Bacteroides ovatus* | 9.02E-06 | 0.00160417 | 0.03 | Intervention |

**Supplementary Table 6.** Reported food intakes that significantly correlated with abundance of bacterial species in participants with CD or UC (highlighted in gray) throughout the duration of the study.

| Food category | *Bacteria species* | Correlation | *p value* | Diagnosis dependent correlation |
|---|---|---|---|---|
| Adverse foods | *Collinsella stercoris* | 0.183509012 | 0.017604064 | CD |
| Adverse foods | *Parabacteroides distasonis* | 0.216207455 | 0.005011379 | CD |
| Adverse foods | *Bifidobacterium longum* | 0.356109274 | 0.000343712 | UC |
| Adverse foods | *Faecalibacterium prausnitzii* | -0.24047139 | 0.017663718 | UC |
| Adverse foods | *Parabacteroides merdae* | 0.223679509 | 0.027636049 | UC |
| Adverse foods\| food & beverages with high sugar | *Eubacterium sp CAG 38* | 0.224008962 | 0.00361236 | CD |
| Adverse foods\| food & beverages with high sugar | *Parabacteroides distasonis* | 0.193573251 | 0.012193904 | CD |
| Adverse foods\| food & beverages with high sugar | *Bifidobacterium longum* | 0.428215985 | 1.21E-05 | UC |
| Adverse foods\| food & beverages with high sugar | *Olsenella scatoligenes* | 0.201165129 | 0.048176276 | UC |
| Adverse foods\| fried vegetable protein | *Anaeromassilibacillus sp An250* | 0.308081409 | 5.11E-05 | CD |
| Adverse foods\| fried vegetable protein | *Eubacterium ventriosum* | 0.212617311 | 0.005804997 | CD |
| Adverse foods\| fried vegetable protein | *Firmicutes bacterium CAG 65* | -0.184680212 | 0.016882441 | CD |
| Adverse foods\| high fat animal protein | *Bacteroides ovatus CAG 22* | 0.164792516 | 0.033323381 | CD |
| Adverse foods\| high fat animal protein | *Bacteroides sp CAG 144* | 0.214840405 | 0.034579769 | UC |
| Adverse foods\| high fat animal protein | *Collinsella intestinalis* | 0.497046405 | 2.23E-07 | UC |
| Adverse foods\| high fat animal protein | *Eubacterium sp CAG 38* | 0.372793634 | 0.000169576 | UC |
| Adverse foods\| high fat animal protein | *Flavonifractor plautii* | 0.291773685 | 0.003734463 | UC |
| Adverse foods\| high fat animal protein | *Parabacteroides merdae* | 0.520360099 | 4.68E-08 | UC |
| Adverse foods\| high fat animal protein | *Roseburia hominis* | 0.295014403 | 0.003351924 | UC |
| Adverse foods\| high fat animal protein | *Ruminococcus sp CAG 177* | 0.56441173 | 1.73E-09 | UC |
| Adverse foods\| lactose | *Flavonifractor plautii* | -0.161917265 | 0.036570104 | CD |
| Adverse foods\| lactose | *Bacteroides stercoris* | 0.335819326 | 0.000771627 | UC |
| Adverse foods\| lactose | *Eubacterium ramulus* | 0.352726784 | 0.00039481 | UC |

| | | | | |
|---|---|---|---|---|
| Adverse foods\| lactose | *Faecalibacterium prausnitzii* | -0.280173327 | 0.005444025 | UC |
| Adverse foods\| lactose | *Flavonifractor plautii* | 0.211156185 | 0.037879296 | UC |
| Adverse foods\| processed & fried animal protein | *Bacteroides ovatus* | -0.18800004 | 0.014975011 | CD |
| Adverse foods\| processed & fried animal protein | *Collinsella intestinalis* | 0.211914877 | 0.005972802 | CD |
| Adverse foods\| processed & fried animal protein | *Eubacterium sp CAG 38* | 0.180068834 | 0.019880718 | CD |
| Adverse foods\| processed & fried animal protein | *Eubacterium ventriosum* | 0.22700947 | 0.003175785 | CD |
| Adverse foods\| processed & fried animal protein | *Alistipes shahii* | 0.230611288 | 0.023054071 | UC |
| Adverse foods\| processed & fried animal protein | *Bacteroides vulgatus* | 0.481662395 | 5.89E-07 | UC |
| Adverse foods\| processed & fried animal protein | *Eubacterium sp CAG 38* | 0.401158863 | 4.66E-05 | UC |
| Adverse foods\| processed & fried animal protein | *Eubacterium ventriosum* | 0.257382953 | 0.01092468 | UC |
| Adverse foods\| processed & fried animal protein | *Ruminococcus sp CAG 177* | 0.265630817 | 0.008546879 | UC |
| Adverse foods\| selected condiments | *Parabacteroides distasonis* | -0.297789288 | 9.29E-05 | CD |
| Adverse foods\| selected condiments | *Parabacteroides distasonis* | -0.200747322 | 0.048652905 | UC |
| Adverse foods\| selected gluten free grains | *Blautia sp CAG 52* | 0.313616604 | 3.68E-05 | CD |
| Adverse foods\| selected gluten free grains | *Faecalibacterium prausnitzii* | -0.183637992 | 0.017523303 | CD |
| Adverse foods\| selected gluten free grains | *Firmicutes bacterium CAG 110* | 0.219113992 | 0.004441635 | CD |
| Adverse foods\| selected gluten free grains | *Bacteroides sp CAG 144* | 0.386002176 | 9.43E-05 | UC |
| Adverse foods\| selected gluten free grains | *Bacteroides vulgatus* | 0.282225086 | 0.005098635 | UC |
| Adverse foods\| selected gluten free grains | *Blautia sp CAG 52* | 0.347459817 | 0.000488422 | UC |
| Adverse foods\| selected gluten free grains | *Eubacterium sp CAG 38* | 0.411218788 | 2.86E-05 | UC |
| Adverse foods\| selected gluten free grains | *Eubacterium ventriosum* | 0.411353044 | 2.84E-05 | UC |
| Adverse foods\| selected gluten free grains | *Firmicutes bacterium CAG 110* | 0.229064403 | 0.024015973 | UC |
| Adverse foods\| selected gluten free grains | *Ruminococcus sp CAG 177* | 0.523528589 | 3.75E-08 | UC |
| Adverse foods\| selected gluten free grains | *Tyzzerella sp* | 0.237126746 | 0.01935555 | UC |
| Adverse foods\| wheat | *Bacteroides xylanisolvens* | 0.244921904 | 0.001422504 | CD |
| Adverse foods\| wheat | *Coprococcus comes* | 0.322700023 | 2.11E-05 | CD |
| Adverse foods\| wheat | *Eubacterium ventriosum* | 0.207058171 | 0.007256376 | CD |
| Adverse foods\| wheat | *Parabacteroides distasonis* | 0.294888165 | 0.000109482 | CD |
| Adverse foods\| wheat | *Bifidobacterium longum* | 0.36782059 | 0.000210169 | UC |

| | | | | |
|---|---|---|---|---|
| Adverse foods\| wheat | *Burkholderiales bacterium 1 1 47* | 0.204120865 | 0.044913535 | UC |
| Adverse foods\| wheat | *Coprococcus comes* | -0.294806242 | 0.003375397 | UC |
| Adverse foods\| wheat | *Faecalibacterium prausnitzii* | -0.323211987 | 0.001241649 | UC |
| Adverse foods\| wheat | *Flavonifractor plautii* | 0.201403052 | 0.047906586 | UC |
| Alcohol | *Firmicutes bacterium CAG 83* | -0.158199254 | 0.041160822 | CD |
| Alcohol | *Alistipes sp CAG 268* | 0.298853477 | 0.002944451 | UC |
| Alcohol | *Bacteroides dorei CAG 222* | 0.252981353 | 0.012416266 | UC |
| Alcohol | *Bacteroides massiliensis* | 0.302393336 | 0.002608756 | UC |
| Alcohol | *Bacteroides sp CAG 144* | 0.24019959 | 0.017796273 | UC |
| Alcohol | *Bacteroides vulgatus* | 0.419261026 | 1.92E-05 | UC |
| Alcohol | *Barnesiella intestinihominis* | 0.48921469 | 3.68E-07 | UC |
| Alcohol | *Eubacterium sp CAG 38* | 0.499966412 | 1.85E-07 | UC |
| Beneficial foods | *Fusicatenibacter saccharivorans* | 0.24176145 | 0.017046115 | UC |
| Beneficial foods | *Ruminococcus torques* | -0.328606978 | 0.001015456 | UC |
| Beneficial foods\| animal protein (lean) | *Alistipes sp HGB5* | 0.184184485 | 0.017184677 | CD |
| Beneficial foods\| animal protein (lean) | *Bacteroides massiliensis* | 0.22351629 | 0.02775264 | UC |
| Beneficial foods\| animal protein (lean) | *Roseburia hominis* | -0.213886532 | 0.035409958 | UC |
| Beneficial foods\| fatty acids MUFAs & Omega-3 | *Bacteroides dorei* | 0.223545437 | 0.003684427 | CD |
| Beneficial foods\| fatty acids MUFAs & Omega-3 | *Faecalibacterium prausnitzii* | 0.178357364 | 0.021105286 | CD |
| Beneficial foods\| fatty acids MUFAs & Omega-3 | *Sutterella parvirubra* | 0.216109724 | 0.005031626 | CD |
| Beneficial foods\| fatty acids MUFAs & Omega-3 | *Bacteroides dorei* | 0.268755576 | 0.007772801 | UC |
| Beneficial foods\| fatty acids MUFAs & Omega-3 | *Fusicatenibacter saccharivorans* | 0.236009888 | 0.019950694 | UC |
| Prebiotics | *Collinsella stercoris* | -0.18334673 | 0.017706136 | CD |
| Prebiotics | *Eubacterium eligens* | 0.199087022 | 0.009899487 | CD |
| Prebiotics | *Parabacteroides distasonis* | -0.179515723 | 0.020269572 | CD |
| Prebiotics | *Roseburia hominis* | 0.164513139 | 0.033627742 | CD |
| Prebiotics | *Alistipes finegoldii* | 0.241758409 | 0.017047549 | UC |
| Prebiotics | *Anaeromassilibacillus sp An250* | 0.304070106 | 0.002462103 | UC |
| Prebiotics | *Bacteroides eggerthii* | -0.215054655 | 0.034395567 | UC |
| Prebiotics | *Firmicutes bacterium CAG 65* | 0.275485926 | 0.006312171 | UC |
| Prebiotics | *Roseburia hominis* | -0.237511001 | 0.019154351 | UC |
| Prebiotics | *Roseburia sp CAG 45* | 0.235455159 | 0.020252093 | UC |
| Prebiotics | *Ruminococcus torques* | -0.208265486 | 0.04064912 | UC |

| | | | | |
|---|---|---|---|---|
| Prebiotics\| fruits | *Bacteroides dorei* | 0.288805704 | 0.000153622 | CD |
| Prebiotics\| fruits | *Bacteroides vulgatus* | -0.212583031 | 0.005813087 | CD |
| Prebiotics\| fruits | *Blautia sp CAG 52* | -0.396647003 | 1.12E-07 | CD |
| Prebiotics\| fruits | *Eubacterium eligens* | 0.177780665 | 0.021532331 | CD |
| Prebiotics\| fruits | *Eubacterium siraeum* | 0.173156253 | 0.025234326 | CD |
| Prebiotics\| fruits | *Alistipes finegoldii* | 0.465194569 | 1.58E-06 | UC |
| Prebiotics\| fruits | *Alistipes sp CAG 268* | 0.365429447 | 0.000232729 | UC |
| Prebiotics\| fruits | *Anaeromassilibacillus sp An250* | 0.343470772 | 0.000572428 | UC |
| Prebiotics\| fruits | *Bacteroides eggerthii* | -0.330514156 | 0.000944943 | UC |
| Prebiotics\| fruits | *Firmicutes bacterium CAG 65* | 0.399579009 | 5.02E-05 | UC |
| Prebiotics\| fruits | *Roseburia sp CAG 45* | 0.345468453 | 0.000528831 | UC |
| Prebiotics\| fruits | *Ruminococcus gnavus* | -0.221903262 | 0.028927469 | UC |
| Prebiotics\| honey | *Eubacterium eligens* | 0.268972119 | 0.000440123 | CD |
| Prebiotics\| honey | *Faecalibacterium prausnitzii* | 0.166610331 | 0.031399497 | CD |
| Prebiotics\| honey | *Bacteroides ovatus* | 0.245097608 | 0.015534709 | UC |
| Prebiotics\| honey | *Eubacterium eligens* | 0.277057561 | 0.006008322 | UC |
| Prebiotics\| honey | *Fusicatenibacter saccharivorans* | 0.224419286 | 0.027112812 | UC |
| Prebiotics\| oats | *Bacteroides dorei CAG 222* | 0.242854005 | 0.016537736 | UC |
| Prebiotics\| oats | *Firmicutes bacterium CAG 65* | -0.23774267 | 0.019033919 | UC |
| Prebiotics\| vegetables | *Anaeromassilibacillus sp An250* | 0.220877473 | 0.029696293 | UC |
| Prebiotics\| vegetables | *Firmicutes bacterium CAG 65* | 0.273451004 | 0.006725767 | UC |
| Prebiotics\| vegetables | *Roseburia hominis* | -0.238390061 | 0.018700824 | UC |
| Prebiotics\| vegetables | *Roseburia sp CAG 45* | 0.218314382 | 0.031693235 | UC |
| Probiotics | *Alistipes sp CAG 268* | 0.238764243 | 0.001887355 | CD |
| Probiotics | *Erysipelatoclostridium ramosum* | 0.238836104 | 0.001881213 | CD |
| Probiotics\| fermented dairy foods | *Clostridium sp CAG 75* | 0.191522393 | 0.013159565 | CD |
| Probiotics\| fermented dairy foods | *Firmicutes bacterium CAG 110* | 0.154161397 | 0.046684834 | CD |
| Probiotics\| fermented dairy foods | *Fusicatenibacter saccharivorans* | 0.195433894 | 0.011372218 | CD |
| Probiotics\| fermented dairy foods | *Bacteroides dorei CAG 222* | -0.257724569 | 0.01081576 | UC |
| Probiotics\| fermented dairy foods | *Fusicatenibacter saccharivorans* | 0.209126156 | 0.039807313 | UC |
| Probiotics\| fermented foods (no dairy) | *Clostridium sp CAG 217* | 0.264825787 | 0.008756976 | UC |
| Probiotics\| fermented foods (no dairy) | *Tyzzerella sp* | -0.316746608 | 0.001572504 | UC |
